# Tissue damage detected by quantitative gradient echo MRI correlates with clinical progression in non-relapsing progressive MS

**DOI:** 10.1101/2021.08.18.21262247

**Authors:** Biao Xiang, Matthew R. Brier, Manasa Kanthamneni, Jie Wen, Abraham Z. Snyder, Dmitriy A. Yablonskiy, Anne H. Cross

## Abstract

**Background:** Imaging biomarkers of progressive MS are needed. Quantitative gradient recalled echo (qGRE) MRI technique allows evaluation of tissue damage associated with microstructural damage in multiple sclerosis (MS).

**Objective:** To evaluate qGRE-derived R2t* as an imaging biomarker of MS disease progression as compared to atrophy and lesion burden.

**Methods:** Twenty-three non-relapsing progressive MS (PMS), twenty-two relapsing-remitting MS (RRMS) and eighteen healthy control participants were imaged with qGRE at 3T. PMS subjects were imaged and neurologically assessed every nine months over five sessions. In each imaging session, lesion burden, atrophy and R2t* in cortical grey matter (GM), deep GM, normal-appearing white matter (NAWM) were measured.

**Results:** R2t* reductions correlated with neurological impairment cross-sectionally and longitudinally. PMS patients with clinically defined disease progression showed significantly faster decrease of R2t* in NAWM and deep GM compared with the clinically stable PMS group. Importantly, tissue damage measured by R2t* outperformed lesion burden and atrophy as a biomarker of progression during the study period.

**Conclusion:** Clinical impairment and progression correlated with accumulating R2t*-defined microstructural tissue damage in deep GM and NAWM. qGRE-derived R2t* is a potential imaging biomarker of MS progression.

## 1. Introduction

Magnetic resonance imaging (MRI) plays an important role in multiple sclerosis (MS) diagnosis and monitoring ^1-3^. Quantification of new or gadolinium enhancing white matter (WM) lesions (WMLs) by means of T1- or T2-weighted clinical images represents disease activity and serves as an end-point for trials of disease-modifying therapies ^4^. However, this approach to disease monitoring applies primarily to relapsing remitting MS (RRMS). Progressive MS (PMS) subtypes [non-relapsing secondary progressive (SP) MS and primary progressive (PP) MS] lack highly sensitive imaging biomarkers of progression due in part to the less inflammatory pathology of PMS ^5, 6^. The best available biomarker of PMS currently is atrophy, especially of grey matter (GM) ^7^. Importantly, atrophy represents the end-result of multiple pathological processes. More specific biomarkers which measure pathology prior to the manifestation of frank atrophy are needed ^8^.

Here, we assess quantitative gradient recalled echo (qGRE) imaging to measure R2t* (=1/T2t*) as a potential biomarker of progression in MS. R2t* is a component of the more commonly reported R2* (=1/T2*) measure and is more specific to tissue microstructure. This study addresses the ability of R2t* to reflect clinical progression and compares the utility of R2t* to atrophy and lesion burden imaging biomarkers. We compare these measures in PMS, RRMS and age-matched healthy control subjects and longitudinally in relation to advancing impairment in PMS patients.

## 2. Materials and methods

### 2.1 Study Participants

Eighteen healthy control (HC), twenty-three PMS (including four PPMS and nineteen SPMS), and twenty-two RRMS subjects were enrolled after providing informed consent. The study was approved by the Institutional Review Board. PMS patients did not have superimposed relapses or gadolinium enhancing lesions within twenty-four months preceding enrollment. RRMS and HC were recruited to reflect the age and sex distribution of the PMS patients. RRMS subjects had no clinical worsening except related to MS attacks. The HC and RRMS subjects were scanned one time at the beginning of the study. PMS subjects were scanned every nine months for five sessions with the goal of identifying imaging markers that correlated best with or predicted clinical progression in PMS patients.

One PMS patient was lost to follow up after the second session, two following the third session, and one following the fourth session. Six imaging sessions were excluded due to severe motion artifacts and six sessions due to technical difficulties. In total, ninety-two sessions (out of 115 possible) in the PMS cohort were used for data analysis.

### 2.2 Clinical testing

Clinical evaluation was performed on the day of MRI exam and included the Expanded Disability Status Scale (EDSS) standardized neurological examination, 25-foot timed walk (25FTW) assessment of gait, nine-hole peg test (9HPT) assessment of bilateral upper extremity function, paced auditory serial addition test (PASAT) and symbol digit modalities test (SDMT) assessments of cognitive function. Clinical assessment was performed by examiners blinded to the imaging results. For data analyses, the 25FTW and 9HPT raw scores were converted to Z-scores according to the Multiple Sclerosis Functional Composite (MSFC) guidelines ^9^.

### 2.3 Definition of clinical progression during the study interval

The PMS subjects were classified as either clinically stable or progressive during the study period based on EDSS, 25FTW and 9HPT scores ^10^. EDSS score increases by one point were defined as progression if the subject’s baseline EDSS was less than 5.5; for baseline EDSS 5.5 or greater, one-half point increases defined progression. For the 25FTW score, ≥ 20% increase or converting from walking to not walking defined progression. For the 9HPT score, increasing by ≥20% or converting from able to unable to complete the test defined progression. Those PMS subjects who did not show progression in EDSS, 25FTW or 9HPT were defined as clinically stable.

### 2.4 Image acquisition

All MRI was performed with a 3.0 Tesla Trio MRI scanner (Siemens, Erlangen, Germany) using a 32-channel phased-array radiofrequency head coil. qGRE data were acquired using three-dimensional multi-gradient-echo sequence with flip angle 30°, TR=50ms, voxel size 1×1×2mm^3^ and acquisition time 12 minutes. 10 gradient echoes, with first echo time TE1=4ms and echo spacing ΔTE=4ms were collected. In addition, fluid-attenuated inversion recovery (FLAIR) images (voxel size 1×1×3mm^3^) and magnetization-prepared rapid gradient-echo (MP-RAGE) (voxel size 1×1×1mm^3^) were acquired for purposes of atlas registration and tissue segmentation.

### 2.5 Image processing and segmentation

R2t* maps were created using the qGRE approach described in a previous study ^11^. In brief, multi-channel data were combined using a previously published algorithm ^12^. Voxel-wise analysis was performed on combined data using the theoretical model of GRE signal relaxation ^13^ and a set of post-processing algorithms that minimize adverse artifacts related to macroscopic magnetic field inhomogeneities ^14^ and physiological fluctuations ^15^ were employed.

Image segmentation was achieved via a two-step process. First, MP-RAGE and FLAIR images were subjected to FreeSurfer analysis (version 6.0) ^16^ and segmentations were reviewed and errors corrected by a trained technician (MK). FreeSurfer does not accurately segment WMLs. Hence, a second approach included application of an intensity-based approach which we recently developed and validated ^17^. For subsequent analyses, voxels were labeled as CSF or GM if both segmentation strategies agreed. Cortical and deep GM were separated based on FreeSurfer segmentation^1^. Normal Appearing White Matter (NAWM) was defined as voxels in which both segmentations returned a WM label. Lesions were defined as voxels identified as lesions by both approaches or where FreeSurfer labeled the voxel as WM and the intensity-based approach labeled the voxel as lesion.

Mean R2t* in cortical grey matter (GM), deep GM, normal appearing white matter 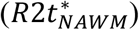, and individual lesions 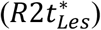 were computed for each subject. In MS patients, to quantity the damage specific to WM lesions, controlling for the damage in NAWM, we defined the Tissue Damage Score (TDS) in each WM lesion as

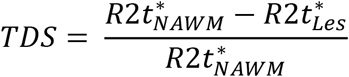

TDS measured in this way quantifies the damage within lesions relative to the NAWM which, in MS patients, may also be abnormal ^17^.

### 2.6 Statistical analysis

Longitudinal analysis of clinical test performance and imaging biomarkers in PMS subjects was conducted via ANOVA with subject as repeated, within-subject factor; the Dunnett test was used to correct for multiple comparison. Baseline clinical and imaging data were compared between subject groups using a one-way ANOVA with group (HC, RRMS, PMS) as factor; the Tukey test was used to correct for multiple comparison.

Spearman rank correlation tests were performed to assess the relationship between imaging biomarkers and clinical test scores. Imaging biomarkers included lesion volume, R2t* measured in cortical and deep GM, NAWM, and TDS. Clinical test performance measures included EDSS, 25FTW, 9HPT, PASAT and SDMT. This cross-sectional analysis used only the first imaging session in the PMS patients. Age and gender were controlled as covariates. False discovery rate was used to correct for multiple comparisons ^18^.

Linear fitting was used to calculate the rate of change of imaging biomarkers in each PMS patient across available visits. Two-sample t-tests were used to compare the rate of change and baseline measurements for stable PMS versus PMS with clinically defined progression.

A major objective of the present work is to isolate imaging biomarkers that predict clinical progression. However, these biomarkers are characterized by high levels of shared variance. Accordingly, to isolate the most predictive biomarkers, LASSO regression was applied to identify the most salient imaging biomarkers of clinical progression ^19^. LASSO is a modification to linear regression wherein regression coefficients (β) values are penalized such that small or redundant β values are forced to zero. Thus, LASSO models are sparse: several β values are large while many are identically 0 indicating those variables do not contribute to the model. Sparsity is controlled by a tuning variable, λ, chosen by cross-validation.

## 3. Results

Demographic information and clinical test performance are summarized in Table 1. Age was well matched between cohorts. The RRMS patients were chosen to match the PMS patients, and thus had an average age of 54.9 years with a range spanning 5 decades. PMS subjects had significantly higher baseline EDSS, 25FTW and 9HPT (dominant and non-dominant) scores compared to RRMS subjects (p<0.001, p<0.01, p<0.001, p<0.001, respectively). The RRMS cohort performed significantly better on SDMT than the PMS cohort (p<0.001). No differences were found in PASAT performance.

**Table 1.** Demographic and clinical information of healthy controls and multiple sclerosis subjects.

|  |  | PMS | RRMS | Healthy Control |
| --- | --- | --- | --- | --- |
| Number of Subjects |  | 23 | 22 | 18 |
| Number of imaging sessions |  | 92 | 22 | 18 |
| mean Age ± SD (years)<br>(range) | | 56.8±8.6<br>(31-75) | 54.9±10.1<br>(22-72) | 48.8±14.5<br>(23-76) |
| Female/Male |  | 15/8 | 18/4 | 13/5 |
| EDSS mean ± SD<br>(range) | | 6.0±1.5<br>(2.5-8.5) | 2.4±1.1<br>(1-5) | N/A |
| 25FTW mean ± SD (second)<br>(range) | | 46.1±63.7<br>(4.2-165.8) | 4.5±1.1<br>(2.7-7.7) | N/A |
| 9HPT mean ± SD<br>(second)<br>(range) | Dominant | 116.1±231.3<br>(18.7-777) | 22.8±6.2<br>(15.2-45.5) | N/A |
| | Non-dominant | 127.4±251.1<br>(21.4-777) | 22.0±3.6<br>(16.3-31.8) | N/A |
| SDMT mean ± SD<br>(range) | | 41.5±14.4<br>(10-63) | 54.7±8.2<br>(37-80) | N/A |
| 3 sec PASAT mean ± SD<br>(range) | | 43.5±13.4<br>(14-60) | 44.9±10.9<br>(27-58) | N/A |

Note: The clinical tests scores of PMS subjects were from session one. Tests scores of subsequent sessions are shown in figure 3.

The PMS group was assessed longitudinally for changes in clinical test scores (Figure 1). Within the PMS group, Visit 5 showed significantly higher (more impaired) scores in EDSS, 25FTW and 9HPT than baseline (p<0.05, p<0.05, p<0.001, respectively). In contrast, cognitive function test scores (PASAT and SDMT) remained stable over the five sessions (p>0.05). Pairwise statistical comparisons for each test and visit are shown in supplemental Table S1. We also compared the performance on clinical tests between the RRMS and PMS group (Figure 1). RRMS compared to the baseline visit of the PMS cohort showed significant differences group-wise in EDSS, 25-foot walk, 9-hole peg test and SDMT (p<0.001, p<0.05, p<0.001, p<0.001, p<0.01, respectively).

**Figure 1.**
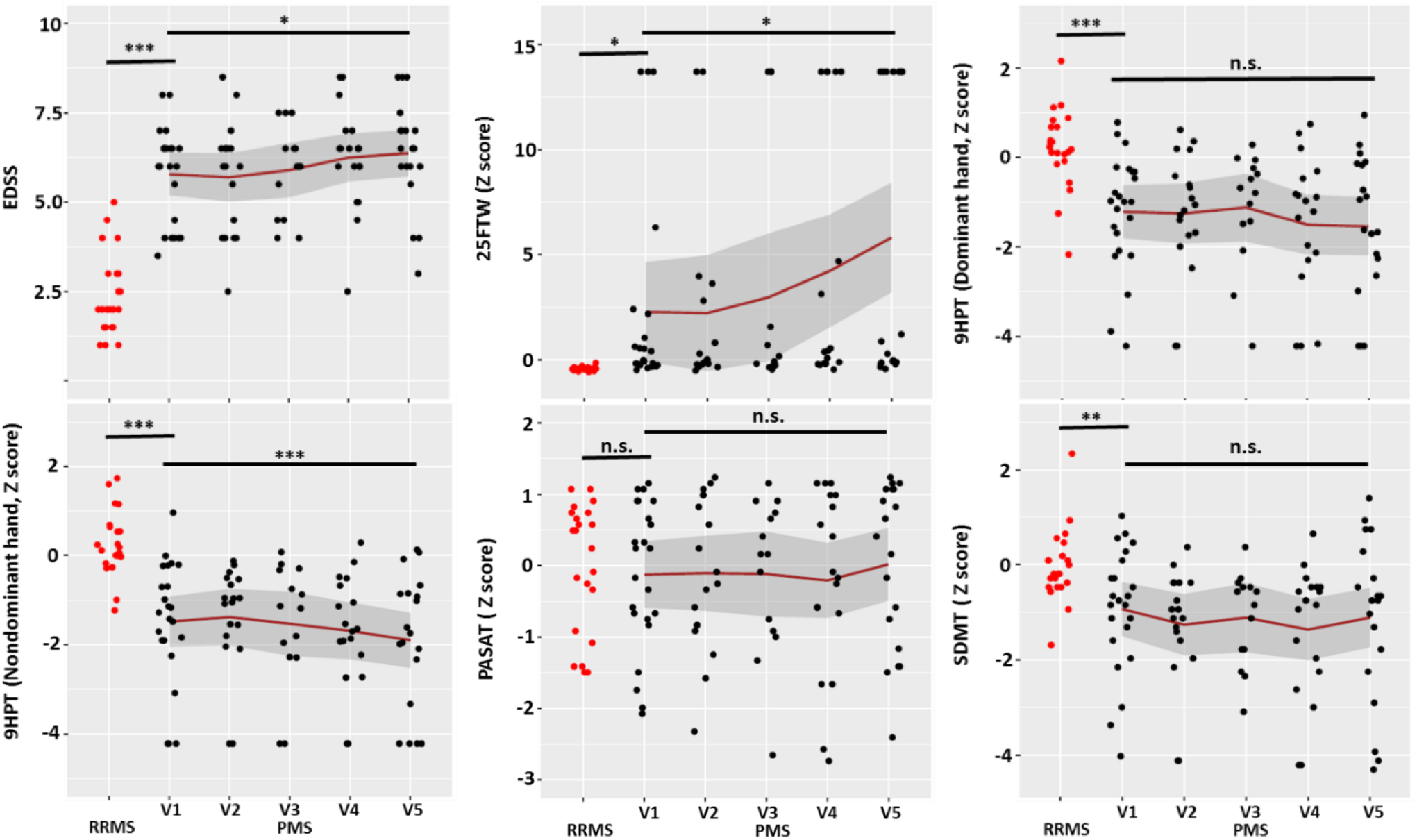
Clinical assessments of the PMS cohort over four years. Compared with the PMS cohort at baseline, the RRMS cohort (red dots) showed significantly better clinical test scores. EDSS, 25-foot walk and 9-hole peg test showed an increasing trend of disability and motor dysfunction for the longitudinal PMS cohort over five imaging sessions, suggesting MS progression within that group. For EDSS, 25FTW and 9HPT (non-dominant hand), there were significant differences between scores in visit five and visit one (p<0.05, p<0.05, p<0.001, respectively). 25FTW, 9HPT, PASAT and SDMT were converted to Z scores. Each dot represents one subject. The gray area represents the standard error. One-way ANOVA with repetitive measurements was used to compare the difference between visit one and each subsequent visit of the PMS group. Dunnett test was used to correct for multiple comparisons. Two-sample t-test was used to compare the RRMS cohort to the first visit of PMS group. *** p < 0.001, ** p <0.01, * p<0.05, n.s. p>0.05.

We next investigated the relation between the R2t* of different brain tissues and clinical test performance. In the cross-sectional analysis of forty-five MS subjects (22 RRMS and 23 PMS), R2t* measurements in NAWM and cortical GM, but not deep GM, correlated significantly with the EDSS, and motor and cognitive test scores (Table 2). The association between most clinical scores and R2t* was stronger in cortical GM compared to NAWM. R2t* in WMLs, quantified relative to NAWM within each subject as the TDS, showed significant correlations with the EDSS and motor-related clinical scores (25FTW, 9HPT), but not cognitive test scores (PASAT, SDMT). Importantly, lesion volume did not show significant correlation with any clinical scores.

**Table 2.** R2t* in the cortical GM, NAWM and lesions correlated with clinical assessments. R2t* measurements in both cortical GM and NAWM correlated with motor and cognitive tests. In addition, R2t* in cortical GM showed stronger correlation with clinical tests than NAWM. Mean lesion tissue damage score (TDS) computed using R2t* measures within lesions showed significant correlations with the EDSS and motor assessments. No significant correlation between lesion volume and clinical scores was found. Spearman rho and p values were computed in R, with age and gender as covariates. All listed p values are after multiple comparison correction using false discovery rate. Statistically significant correlations are highlighted in color.

|  | R2t* (Cortical GM)/s<sup>-1</sup> |  | R2t* (Deep GM)/s<sup>-1</sup> |  | R2t* (NAWM)/s<sup>-1</sup> |  | Mean Lesion TDS |  | Lesion Volume/mm<sup>3</sup> |  |
| --- | --- | --- | --- | --- | --- | --- | --- | --- | --- | --- |
|  | rho | p | rho | p | rho | p | rho | p | rho | p |
| EDSS | -0.722 | p<0.001 | -0.153 | 0.223 | -0.498 | p<0.001 | 0.431 | p<0.001 | 0.199 | 0.091 |
| 25FTW | -0.748 | p<0.001 | -0.246 | 0.034 | -0.531 | p<0.001 | 0.481 | p<0.001 | 0.141 | 0.254 |
| 9HPT (Dominant) | 0.453 | p<0.001 | -0.041 | 0.722 | 0.330 | 0.003 | -0.288 | 0.011 | -0.058 | 0.645 |
| 9HPT (NonDominant) | 0.562 | p<0.001 | -0.028 | 0.786 | 0.422 | p<0.001 | -0.339 | 0.002 | -0.119 | 0.315 |
| PASAT | 0.296 | 0.009 | -0.119 | 0.315 | 0.335 | 0.003 | 0.087 | 0.481 | -0.201 | 0.091 |
| SDMT | 0.448 | p<0.001 | 0.051 | 0.702 | 0.390 | p<0.001 | -0.213 | 0.067 | -0.128 | 0.302 |

R2t* was measured in cortical and deep GM, NAWM and lesions (as TDS) in the RRMS and PMS groups cross-sectionally and the PMS group longitudinally (Figure 2). At baseline, cortical GM R2t* was lower in the PMS cohort compared to RRMS and control group (p<0.05). Lesion TDS showed larger (worse) values in PMS compared to RRMS. In PMS, both cortical and deep GM R2t* declined over the five imaging sessions. R2t* in NAWM did not change across visits, However, the data exhibited increasing variability at later visits, suggesting that R2t* in NAWM declines more in some subjects than others. Pairwise comparisons for each measure and visit are shown in the supporting information (Table S2).

**Figure 2.**
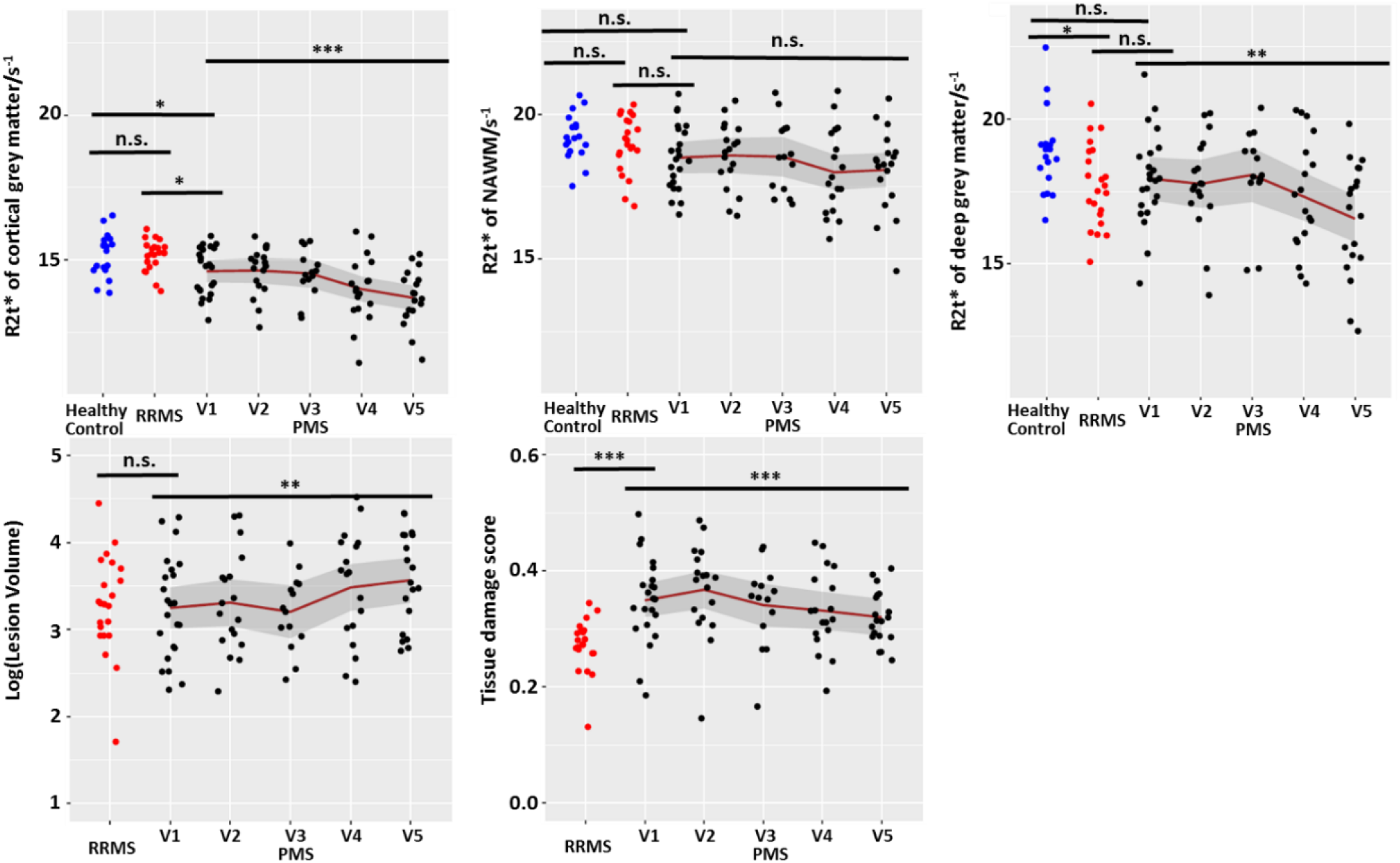
Longitudinal R2t*, lesion volume and tissue damage score measurements for non-relapsing PMS subjects compared with healthy control and RRMS measurements. The longitudinal PMS cohort showed significant decreasing of R2t* (greater microstructural tissue damage) in cortical GM and deep GM over five imaging sessions, performed nine months apart. R2t* of cortical GM and deep GM in visit five were significantly different from R2t* in visit one (p<0.001 and p<0.01, respectively). The R2t* measurements in GM of PMS subjects at baseline were significantly lower than healthy control and RRMS cohorts (p<0.05). Lesion volume of PMS subjects increased over four years. Specifically, lesion volume in visit five was significantly higher than the lesion volume in visit one (p<0.01). Tissue damage score declined over the last four visits, which is likely due to the decrease of R2t* in NAWM, making the differences between R2t* in NAWM and lesion smaller. The line represents the mean value and the gray area represents the standard error. One-way ANOVA with repetitive measurements was used to compare the difference between visit one and each subsequent visit of the PMS group. Dunnett test was used to correct for multiple comparisons. One-way ANOVA was used to compare the healthy control, RRMS and baseline visit one of the PMS group. Tukey test was used to correct for multiple comparisons. *** p < 0.001, ** p <0.01, * p<0.05, n.s p>0.05.

To determine if imaging biomarkers distinguished stable (n=10) vs. clinically progressing (n=13) non-relapsing PMS patients, we calculated the rate of change in R2t*, lesion volume, and atrophy. Two-sample t-tests were used to compare the rates of change between the two groups. The baseline measurements of lesion volume, atrophy and R2t* were not statistically different between groups (Figure 3). PMS subjects with clinically defined progression showed faster decline of R2t* in NAWM and deep grey matter. These statistical tests were significant considered in isolation but not significant when adjusted for multiple comparisons. This result suggests that patients with clinical progression, while not different from stable subjects at baseline, had more rapid tissue damage accumulation in NAWM and deep GM over the study period, detected by R2t* but not by volume loss.

**Figure 3.**
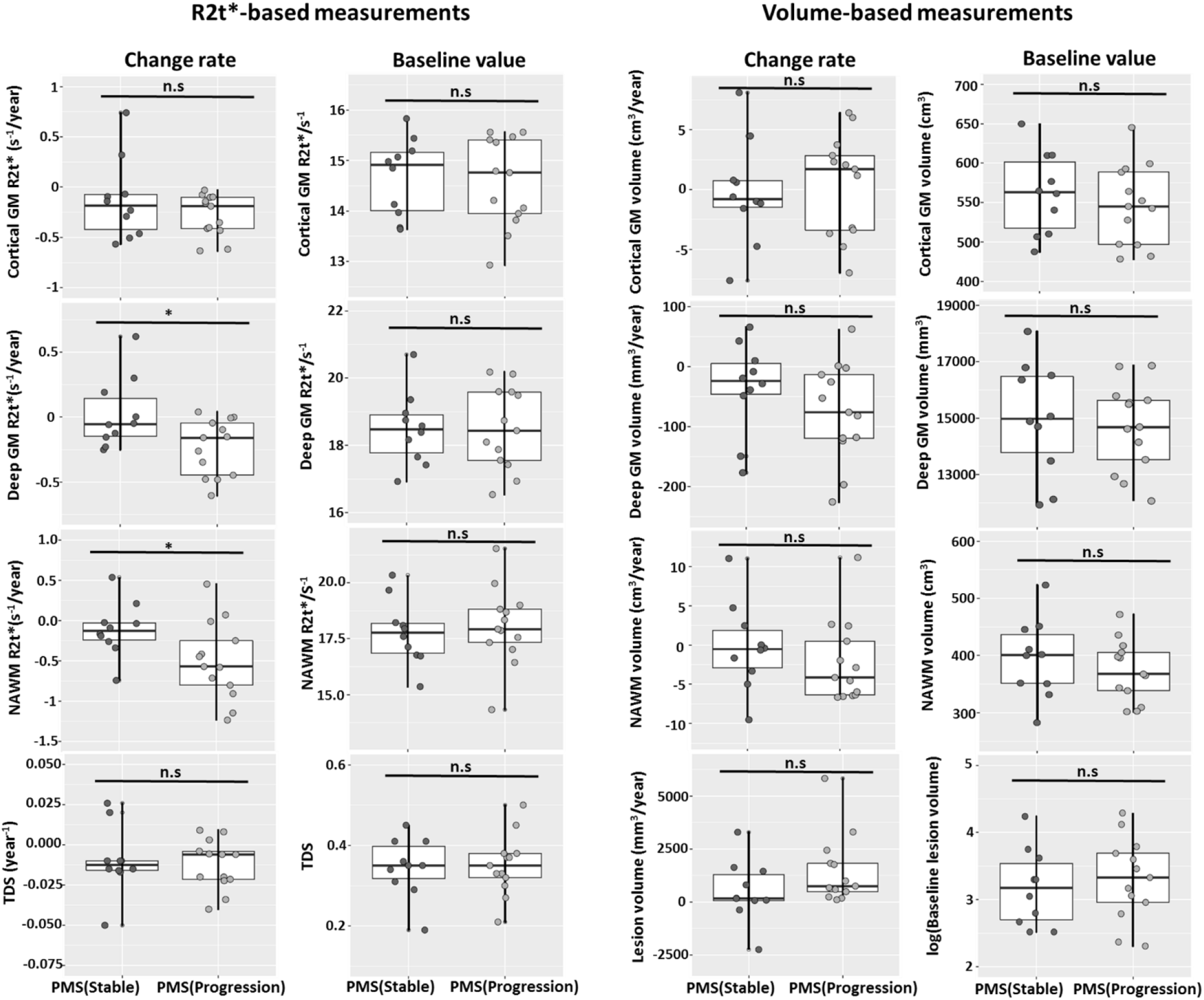
Rate of R2t* decline in NAWM and deep grey matter was faster in PMS subjects with clinically defined progression than in stable PMS subjects. Thirteen PMS subjects showed clinically defined progression, whereas ten subjects were stable on clinical tests. Each dot represents one subject. Each row corresponds to an imaging biomarker. The first and third columns of graphs show rate of change and the second and fourth columns show the baseline values. Rate of change of R2t* in deep grey matter and NAWM were significantly faster in PMS with clinical progression, whereas rate of change in lesion tissue damage score, lesion volume, NAWM volume and deep GM atrophy were nominally greater in the PMS group with clinical progression group than the stable group, but did not reach statistical significance. * p<0.05, n.s p>0.05.

Logistic LASSO was used to identify the unique contribution of changes in R2t* in NAWM and deep GM to the prediction of clinical progression. Predictors included in the model were initial and rate of change in R2t*, lesion volume, and atrophy (Table 3). Imaging data were standardized as Z-scores, resulting in regression coefficients (β values) that are comparable across variables. Baseline imaging values were not strongly associated with later progression. However, change in deep GM and NAWM R2t* were associated with clinical progression. The largest β values were observed in the rate of change in R2t* of deep GM, followed by NAWM and lesion volume. This result is compatible with R2t* of deep GM and NAWM contributing additional predictive information apart from atrophy and lesion burden.

**Table 3:**
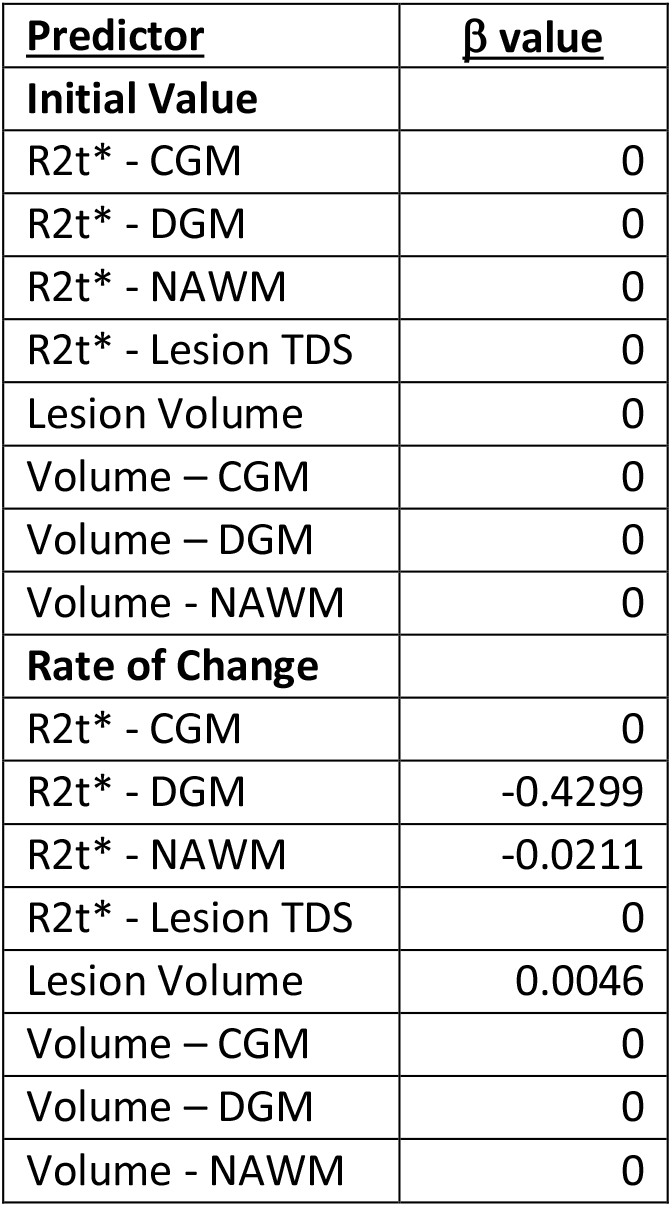
Imaging variables that correlate with clinical progression. LASSO logistic regression was fit with the 10 imaging parameters as predictors and the group (clinically stable PMS vs. clinical progression PMS) as outcome. Input variables were normalized to Z scores such that values exist on the same scale. λ=0.1375, the tuning parameter, was optimized by cross validation. CGM = cortical grey matter; DGM = deep grey matter; NAWM = normal appearing white matter; TDS= tissue damage score.

## 4. Discussion

MRI has significantly contributed to our understanding of MS pathology. WMLs, detected on gadolinium enhanced T1- and T2-weighted sequences, reliably correlate with clinical relapses in RRMS ^4^. Based on this, new or enhancing lesions are included as outcome measures in ongoing clinical trials and represent disease activity potentially due to suboptimal treatment of RRMS ^20^. However, no currently available imaging biomarker, apart from atrophy, similarly correlates with progression in PMS. Because atrophy occurs subsequent to damage, identification of progression by atrophy is possible only in retrospect. This has, at least partially, slowed down development of treatments of PMS. The current results demonstrate that R2t* strongly correlates with clinical impairment more closely than atrophy, appearing to concurrently reflect clinical progression.

qGRE quantitatively measures R2t* (=1/T2t*) ^13^, which is a sub-component of total R2* GRE signal decay (R2*=1/T2*) ^11^. Previous studies showed that quantitative determination of R2* differentiates RRMS patients from healthy controls and correlates with neurological impairment ^12, 21, 22^. R2t* is theoretically more specific to tissue microstructure than total R2* because it measures a part of the GRE signal decay (transverse relaxation rate) that depends strongly on the local cellular-matrix environment of water molecules (a major source of MRI signal). R2* is affected by macroscopic field inhomogeneities, physiological fluctuations and the vascular-related blood oxygen level-dependent (BOLD) effect whereas R2t* is less affected ^13-15^. Thus, R2t* is more closely related to intrinsic tissue integrity than R2* and is a more biologically relevant biomarker. Prior results also showed significant associations between CNS R2t* reduction and MS clinical severity ^23, 24^. Importantly, the present results expand those findings to non-relapsing clinical progression examined longitudinally and demonstrate that R2t*-based measures outperform atrophy- and lesion-based measures as clinical biomarkers.

Histopathological studies have demonstrated that MS tissue damage is complex, involving not only demyelination but also axonal injury and transections, and gliosis in WMLs, GM and NAWM ^25^. Progressive MS is thought to be due to continuing tissue injury mediated by multiple mechanisms beyond inflammatory activity ^26^. We assayed intrinsic tissue damage using R2t* as a marker of pathology in patients with RRMS and PMS. We showed that PMS patients had greater tissue injury (reduced R2t*) compared to similarly aged RRMS patients and that this injury correlated closely with clinical test performance. Consistent with ongoing damage, we found longitudinal reductions in R2t* across much of the brain in PMS patients. PMS patients were further stratified as progressing or stable over the study period. Almost all PMS patients demonstrated at least mildly worsened (but not always statistically significant) clinical test performance across the study period. Importantly, patients with confirmed progression, indicating significantly worsened impairment, had the most dramatic declines in tissue integrity in deep GM and NAWM. The association of deep GM atrophy with MS-related disability, specifically in progressive disease, is well documented ^27, 28^ but tissue atrophy is likely the end result of multiple pathologies. The present results suggest that tissue integrity as measured by R2t* reflects neuropathology not yet manifesting as atrophy.

This study has a few limitations. The cohort was relatively small (23 PMS patients). Nevertheless, we detected differential changes in those who progressed versus those who did not, suggesting that R2t* might serve as a surrogate for progression in early phase clinical trials in PMS. The progressive patients were followed every nine months for five visits, a short time compared to the time-course of PMS. Some patients may have clinically progressed but remained below the detection threshold. In addition, learning effects might have masked cognitive decrements on repeated testing. We were able to match RRMS and PMS cohorts by age but matching the two subtypes by both age and impairment was not possible due to the near-definitional requirement for PMS to have more significant clinical impairment. The histological basis of R2t* signal changes in the deep GM is uncertain. Measured R2t* reflects the combined effects of microstructural tissue damage and non-heme iron deposition. This question will be a subject of our future studies employing a method for separation of cellular and iron contributions to GRE signal recently developed by our group ^29^.

## 5. Conclusion

We measured qGRE R2t* as a biomarker for tissue microstructural damage in brains of RRMS and PMS patients. We examined longitudinal changes in R2t* in non-relapsing PMS patients with and without clinically confirmed progression over the study period. We found that decreased R2t* in deep GM and NAWM correlated with clinical progression. This finding supports the potential of R2t* as a quantitative imaging biomarker of ongoing MS progression.

## Data Availability

The data that support the findings of this study are available from the corresponding author, Anne H. Cross, upon reasonable request.

## Supporting Information

**Table S1.**
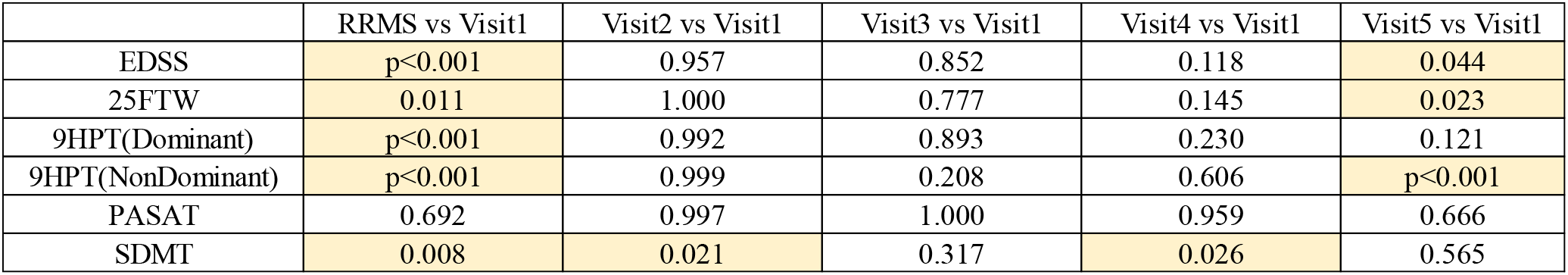
p values of group comparisons for clinical assessments between the RRMS and PMS groups and within the PMS group. Significant difference was observed between the RRMS and PMS groups for EDSS, 25FTW, 9HPT and SDMT. EDSS, 25FTW and 9HPT of visit five of the PMS group was significantly different than visit one of the PMS group. One way ANOVA with repetitive measurements was used to compare the difference between visit one and each subsequent visit of the PMS group. Dunnett test was used to correct for multiple comparison. Two-sample t-test was used to compare the RRMS and the first visit of PMS group. Statistically significant correlations are highlighted in color.

|  | RRMS vs Visit1 | Visit2 vs Visit1 | Visit3 vs Visit1 | Visit4 vs Visit1 | Visit5 vs Visit1 |
| --- | --- | --- | --- | --- | --- |
| EDSS | p<0.001 | 0.957 | 0.852 | 0.118 | 0.044 |
| 25FTW | 0.011 | 1.000 | 0.777 | 0.145 | 0.023 |
| 9HPT(Dominant) | p<0.001 | 0.992 | 0.893 | 0.230 | 0.121 |
| 9HPT(NonDominant) | p<0.001 | 0.999 | 0.208 | 0.606 | p<0.001 |
| PASAT | 0.692 | 0.997 | 1.000 | 0.959 | 0.666 |
| SDMT | 0.008 | 0.021 | 0.317 | 0.026 | 0.565 |

**Table S2.**
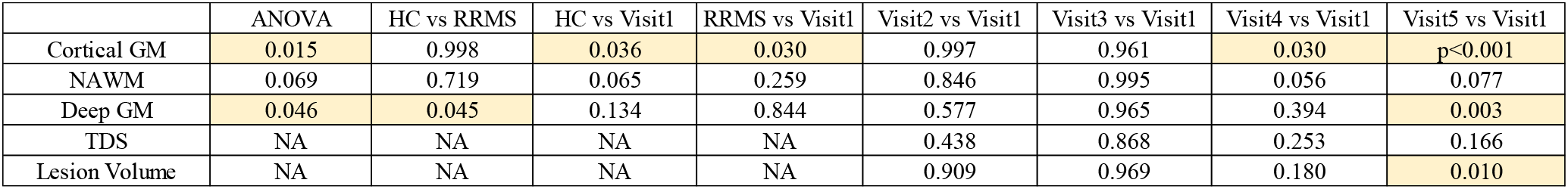
p values of group comparisons for MRI measurements between the healthy control, RRMS and PMS groups, and within the PMS group. One way ANOVA with repetitive measurements was used to compare the difference between visit one and each subsequent visit of the PMS group. Dunnett test was used to correct for multiple comparison. One way ANOVA was used to compare the healthy control, RRMS and visit one of the PMS group. Tukey test was used to correct for multiple comparison. Statistically significant correlations are highlighted in color.

|  | ANOVA | HC vs RRMS | HC vs Visit1 | RRMS vs Visit1 | Visit2 vs Visit1 | Visit3 vs Visit1 | Visit4 vs Visit1 | Visit5 vs Visit1 |
| --- | --- | --- | --- | --- | --- | --- | --- | --- |
| Cortical GM | 0.015 | 0.998 | 0.036 | 0.030 | 0.997 | 0.961 | 0.030 | p<0.001 |
| NAWM | 0.069 | 0.719 | 0.065 | 0.259 | 0.846 | 0.995 | 0.056 | 0.077 |
| Deep GM | 0.046 | 0.045 | 0.134 | 0.844 | 0.577 | 0.965 | 0.394 | 0.003 |
| TDS | NA | NA | NA | NA | 0.438 | 0.868 | 0.253 | 0.166 |
| Lesion Volume | NA | NA | NA | NA | 0.909 | 0.969 | 0.180 | 0.010 |

## Footnotes

1 Deep GM included the basal ganglia (caudate, putamen, globus pallidus), and thalamic nuclei.

